# Phylogenetic-based inference reveals distinct transmission dynamics of SARS-CoV-2 variant of concern Gamma and lineage P.2 in Brazil

**DOI:** 10.1101/2021.10.24.21265116

**Authors:** Tiago Graf, Gonzalo Bello, Felipe Gomes Naveca, Marcelo Gomes, Vanessa Leiko Oikawa Cardoso, Alexandre Freitas da Silva, Filipe Zimmer Dezordi, Mirleide Cordeiro dos Santos, Katia Correa de Oliveira Santos, Erika Lopes Rocha Batista, Alessandro Leonardo alvares Magalhaes, Fernando Vinhal, Brazilian Ministry of Health COVID-19 Genomic Surveillance Network, Fabio Miyajima, Helisson Faoro, Ricardo Khouri, Gabriel Luz Wallau, Edson Delatorre, Marilda Mendonca Siqueira, Paola Cristina Resende

**Affiliations:** Instituto Goncalo Moniz, Fundacao Oswaldo Cruz, Salvador, Brazil; Laboratorio de AIDS e Imunologia Molecular, Instituto Oswaldo Cruz, FIOCRUZ, Rio de Janeiro, Brazil; Laboratorio de Ecologia de Doencas Transmissiveis na Amazônia (EDTA), Leônidas e Maria Deane Institute, Fiocruz; Grupo de Metodos Analiticos em Vigilancia Epidemiologica, Programa de Computacao Cientifica (PROCC), Fiocruz, Rio de Janeiro, Brazil; Instituto Aggeu Magalhaes, Fundacao Oswaldo Cruz, Recife, Pernambuco, Brazil; Instituto Evandro Chagas, Belem, Para; Instituto Adolfo Lutz, Sao Paulo; Secretaria de Saude de Aparecida de Goiania, Goias, Brazil; Lab HLAGYN - Goiania, Goias, Brazil; Brazilian Ministry of Health COVID-19 Genomic Surveillance Network, Av. Brasil, 4365 - Manguinhos, Rio de Janeiro - CEP: 21040-900; Fundacao Oswaldo Cruz - Fiocruz Ceara; Instituto Carlos Chagas (ICC), Fiocruz-PR; Departamento de Biologia. Centro de Ciencias Exatas, Naturais e da Saude, Universidade Federal do Espirito Santo, Alegre, Brazil; Laboratory of Respiratory Viruses and Measles, Oswaldo Cruz Institute (IOC), Oswaldo Cruz Foundation (FIOCRUZ), Rio de Janeiro, RJ, Brazil

**Author notes:** **Corresponding authors:** Tiago Graf, Gonzalo Bello. These authors contributed equally to this work (Tiago Graf, Gonzalo Bello). List of authors and affiliations (Appendix 1).

## Abstract

The COVID-19 epidemic in Brazil experienced two major country-wide lineage replacements, the first driven by the lineage P.2, formerly classified as variant of interest (VOI) Zeta in late 2020 and the second by the variant of concern (VOC) Gamma in early 2021. To better understand how these SARS-CoV-2 lineage turnovers occurred in Brazil, we analyzed 11,724 high-quality SARS-CoV-2 whole genomes of samples collected in different country regions between September 2020 and April 2021. Our findings indicate that the spatial dispersion of both variants in Brazil was driven by short and long-distance viral transmission. The lineage P.2 harboring Spike mutation E484K probably emerged around late July 2020 in the Rio de Janeiro (RJ) state, which contributed with most (∼50%) inter-state viral disseminations, and only became locally established in most Brazilian states by October 2020. The VOC Gamma probably arose in November 2020 in the Amazonas (AM) state, which was responsible for 60-70% of the inter-state viral dissemination, and the earliest timing of community transmission of this VOC in many Brazilian states was already traced to December 2020. We estimate that variant Gamma was 1.56-3.06 more transmissible than variant P.2 co-circulating in RJ and that the median effective reproductive number (Re) of Gamma in RJ and SP states (Re = 1.59-1.91) was lower than in AM (Re = 3.55). In summary, although the epicenter of the lineage P.2 dissemination in Brazil was the heavily interconnected Southeastern region, it displayed a slower rate of spatial spread than the VOC Gamma originated in the more isolated Northern Brazilian region. Our findings also support that the VOC Gamma was more transmissible than lineage P.2, although the viral Re of the VOC varied according to the geographic context.

## Introduction

The COVID-19 epidemic in Brazil during 2020 was mostly driven by SARS-CoV-2 lineages B.1.1.28 and B.1.1.33 in most country regions, and by lineages B.1.1 and B.1.195 at some specific states ^1-3^. In late 2020, two descendant sub-lineages of B.1.1.28 emerged in Brazil and replaced dominant lineages across the country. The first lineage turnover observed during late 2020 was associated with the emergence and dissemination of the lineage P.2, formerly defined Variant of Interest (VOI) Zeta, which harbors mutation E484K in the Spike protein ^4^. The lineage P.2 was first detected in the Rio de Janeiro (RJ) state in October 2020 and became the most prevalent lineage in several Brazilian states from November 2020 to January 2021 ^4-6^. The second country-wide lineage turnover event took place after the emergence of the Variant of Concern (VOC) Gamma in the Amazonas (AM) state in November 2020 ^3,7^. The VOC Gamma harbors multiple mutations in the Spike protein and becomes the dominant lineage in all Brazilian regions by early 2021 ^6,8-11^.

Phylodynamic reconstruction of the spatiotemporal spread pattern of lineages B.1.1.28 and B.1.1.33 supports that densely populated and well-connected urban centers in the Southeastern region were the main sources of inter-regional SARS-CoV-2 spread during the first epidemic wave in 2020 in Brazil ^2^. Mathematical modeling also supports that São Paulo (SP) and RJ were the main superspreader cities of SARS-CoV-2 during the first months of the pandemic in Brazil ^12^. Even though lineage P.2 and VOC Gamma spread widely through Brazil since late 2020, the routes of regional dissemination and the onset date of community transmission across different states remains unclear. Furthermore, despite previous studies estimated that the effective reproductive number (Re) of the VOC Gamma was nearly two times higher than non-VOC co-circulating in the AM state ^3,7,13^, the Re of this VOC in other states and its relative transmissibility concerning the lineage P.2 has not been addressed yet.

Here, we used a large dataset of SARS-CoV-2 positive samples sequenced by the Brazilian Ministry of Health COVID-19 Genomic Surveillance Network (see Methods for details). Genomes retrieved from patients of all Brazilian states sampled between September 2020 and April 2021 were analyzed alongside publicly available SARS-CoV-2 sequences sampled throughout Brazil in the same period. Our findings show the changing prevalence of lineage P.2 and VOC Gamma in different country regions, reconstruct the pathways of inter-state spread, estimate the onset date of community transmission in such locations and compare the relative transmissibility of both co-circulating variants.

## Results

### Genomic surveillance of SARS-CoV-2 in Brazil: September 2020 to March 2021

The COVID-19 epidemic in Brazil displayed two distinct phases over the period of investigation: a decreasing phase from around 900 deaths/day in early September to 300 deaths/day in mid-November 2020; followed by an increasing phase up to 3,000 deaths/day in late March 2021 (**Fig 1a**). A total of 11,724 high quality (<5% N) Brazilian SARS-CoV whole-genomes (29 kb) sampled between September 01^st^, 2020 and April 16^th^, 2021 were analyzed, including 5,472 generated by our collaborative genomic surveillance network (**Fig1b**) and 6,252 obtained from the EpiCoV database in GISAID (https://www.gisaid.org/), covering all Brazilian states (**Supplementary Table 1**). Based on this sampling, our genomic analysis revealed a changing pattern of SARS-CoV-2 lineage frequencies in the studied period with a high overall prevalence of lineages B.1.1.28 and B.1.1.33 during the decreasing phase, a subsequent predominance of lineage P.2 from December 2020 to January 2021, and the dominance of the VOC Gamma in February and March 2021 (**Fig 1c**). In addition, it is interesting to note that the overall proportion of local variants harbouring the mutation Spike:E484K, which comprises the VOC Gamma and the lineages P.2, N.9, N.10, sharply increased in Brazil from <1% in September 2020 to >90% in March 2021 (**Fig 1d**).

**Figure 1.**
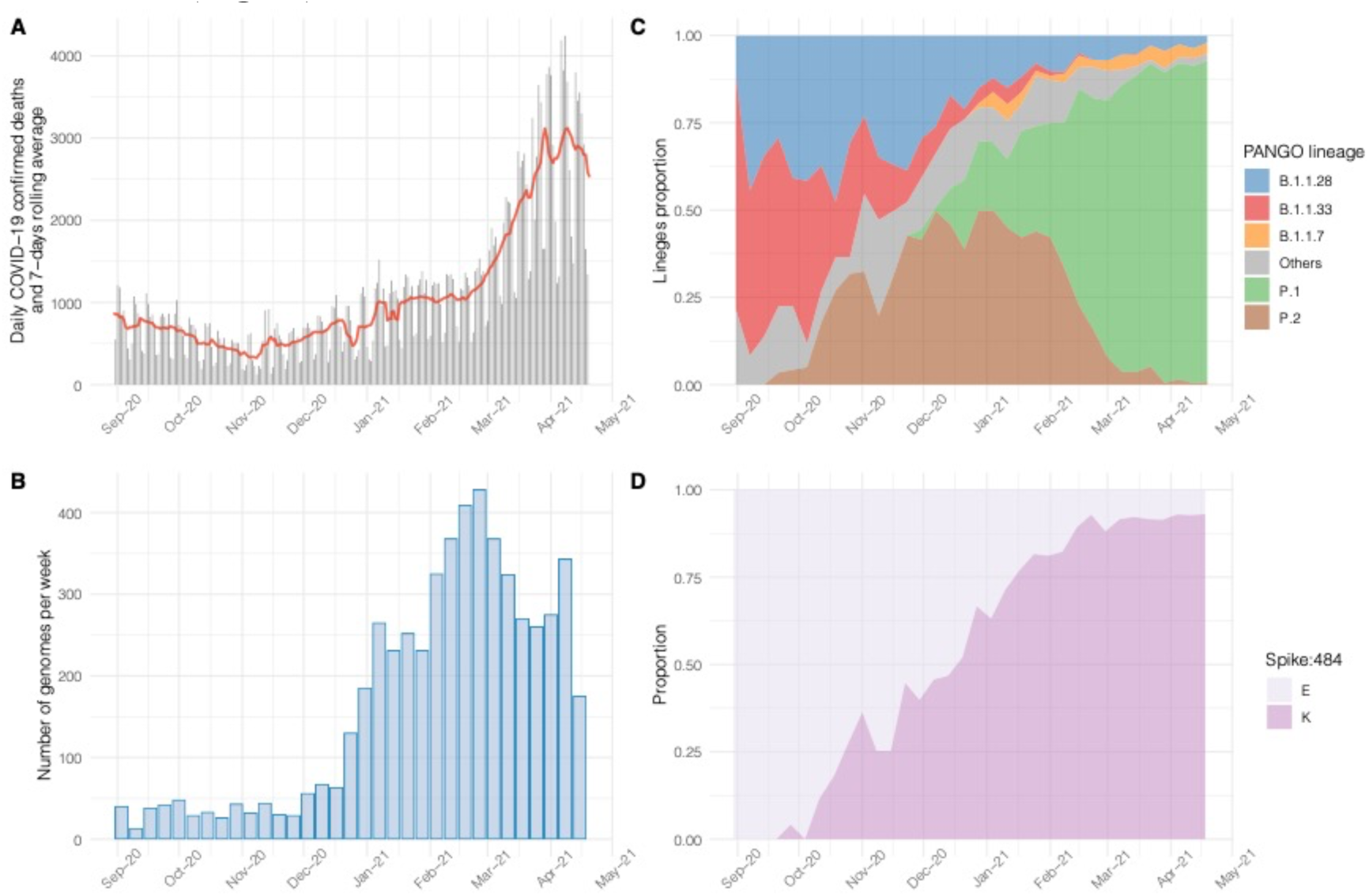
Genomic epidemiology of COVID-19 in Brazil from September 2020 to March 2021. a) Daily number of deaths due to COVID-19 and 7-days rolling average (line). b) Weekly number of genomes generated in this study. c) Proportion of lineages circulating in Brazil in the study period. d) Proportion of variants harboring Spike:484E/K in Brazil in the study period.

### Timing the origin of variants Gamma and P.2 in Brazil

To estimate the precise onset date of the lineage P.2 and the VOC Gamma, we first excluded all P.1 or P.2 sequences that did not display the full set of variant-defining mutations and those recently classified as Gamma-like ^14^. Next, we assessed the temporal signal of the P.2 and Gamma datasets and excluded several outliers, including some of the sequences with the earliest recorded sampling dates (**Fig 2a**). The leaf-dating Bayesian molecular clock method confirmed that the earliest Gamma sequence recovered in November 2020 and three P.2 genomes sampled between April and August 2020 clearly violate the assumption of a molecular clock as displayed median expected collection dates much more recent than their recorded sampling dates (**Fig 2b**). This quality control process resulted in two final datasets of 1,357 P.2 and 2,477 Gamma Brazilian sequences sampled between 25^th^ September 2020 - 26^th^ March 2021 and 03^rd^ December 2020 - 16^th^ April 2021, respectively, mostly (>70%) generated by our collaborative genomic surveillance network. Finally, to estimate the time of the most recent common ancestor (T_MRCA_) of each variant we generated spatiotemporal representative subsets containing 219 P.2 and 191 Gamma sequences. Bayesian molecular clock analyses traced the T_MRCA_ of the lineage P.2 to 28^th^ July 2020 (95% HPD: 11^th^ June-3^rd^ September 2020) and of that of the VOC Gamma to 30^th^ November 2020 (95% HPD: 14^th^ November-3^rd^ December 2020) (**Fig 2c**).

**Figure 2.**
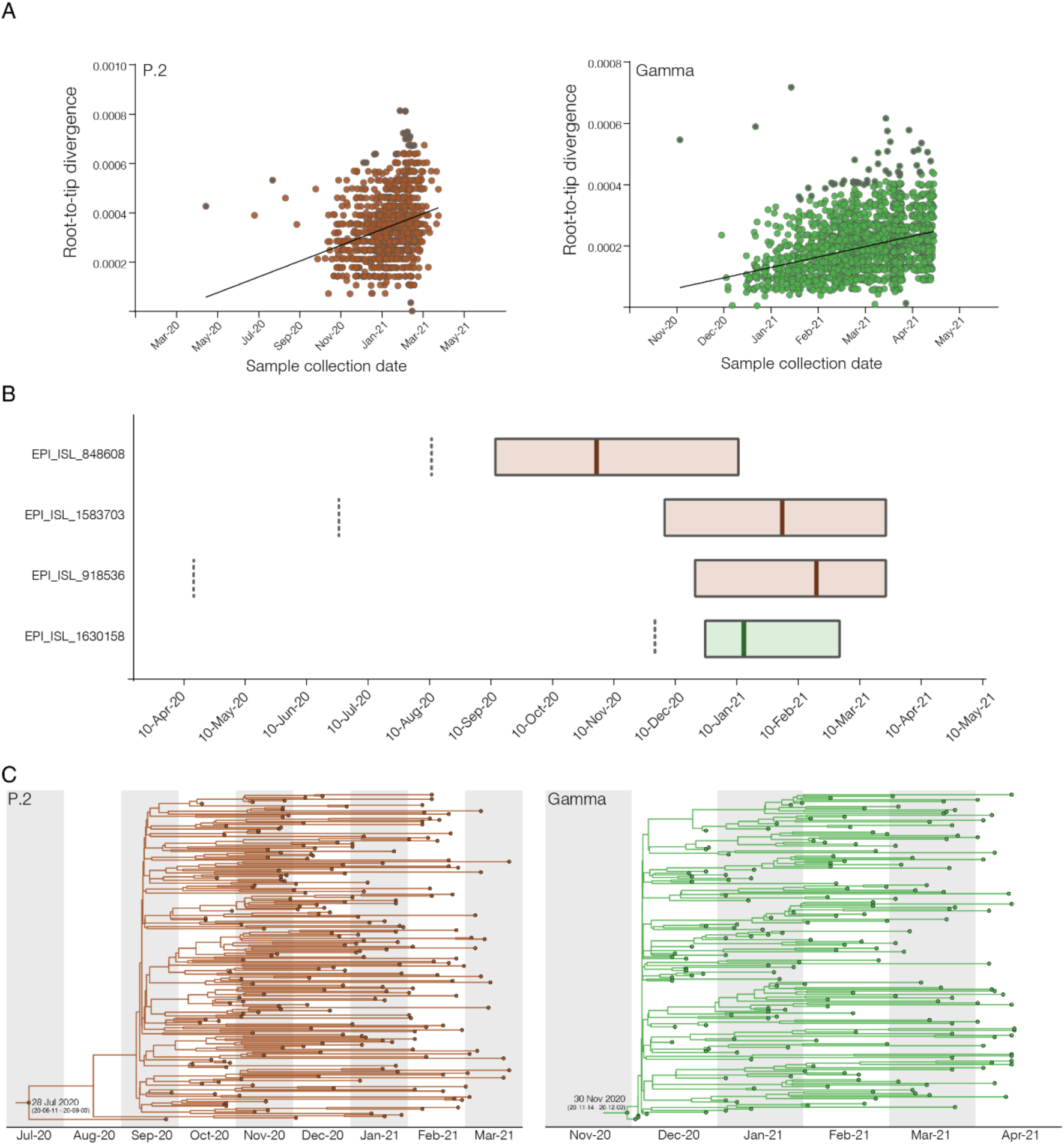
Temporal analysis of the P.2 and Gamma sequence datasets. a) Plots of the root-to-tip divergence against dates of sample collection for P.2 and Gamma datasets. Outliers excluded from the further analysis were colored gray. b) Leaf-dating Bayesian molecular clock of the four sequences excluded due to probably wrong collection date annotation. Reported collection dates are indicated by the dotted lines. Boxplots represent the median and the 95% highest posterior density interval of the estimated collection dates. c) Time-calibrated maximum clade credibility tree for P.2 and Gamma lineages spatial-temporal representative datasets. The median estimated onset dates (with 95% highest posterior density interval in parentheses) for each clade are shown on the root

### Inter-state spread and domestic dissemination of variants Gamma and P.2 in Brazil

To describe the pathways that shaped the country-wide dissemination of the lineage P.2 and the VOC Gamma in Brazil, we performed a discrete Bayesian phylogeographic reconstruction. Bayesian analyses support that the lineage P.2 most probably (*PSP* = 0.71) arose in RJ (Southeastern region). This state was also the main epicenter of the regional spread of this lineage in Brazil, followed by the Southern states of Santa Catarina (SC) and Rio Grande do Sul (RS) and the Southeastern state of SP (**Fig 3a and Supplementary Video 1**). RJ contributed with ∼50% of all inter-state disseminations of lineage P.2 between October 2020 and February 2021, while the four major spreading states (RJ, SP, SC and RS) combined accounted for >90% of all inter-state viral disseminations in that period (**Fig 3b**). The relative contribution of the secondary hubs SC, RS and SP to the overall dissemination of P.2 remained roughly constant between mid-October 2020 and late February 2021. The P.2 migrations from RJ to other states started in September 2020 but were mostly distributed between October 2020 and January 2021 (**Fig 3c**). The earliest onset date of community transmission of the lineage P.2 was traced to October 2020 in most Brazilian states **(Fig 3d**).

**Figure 3.**
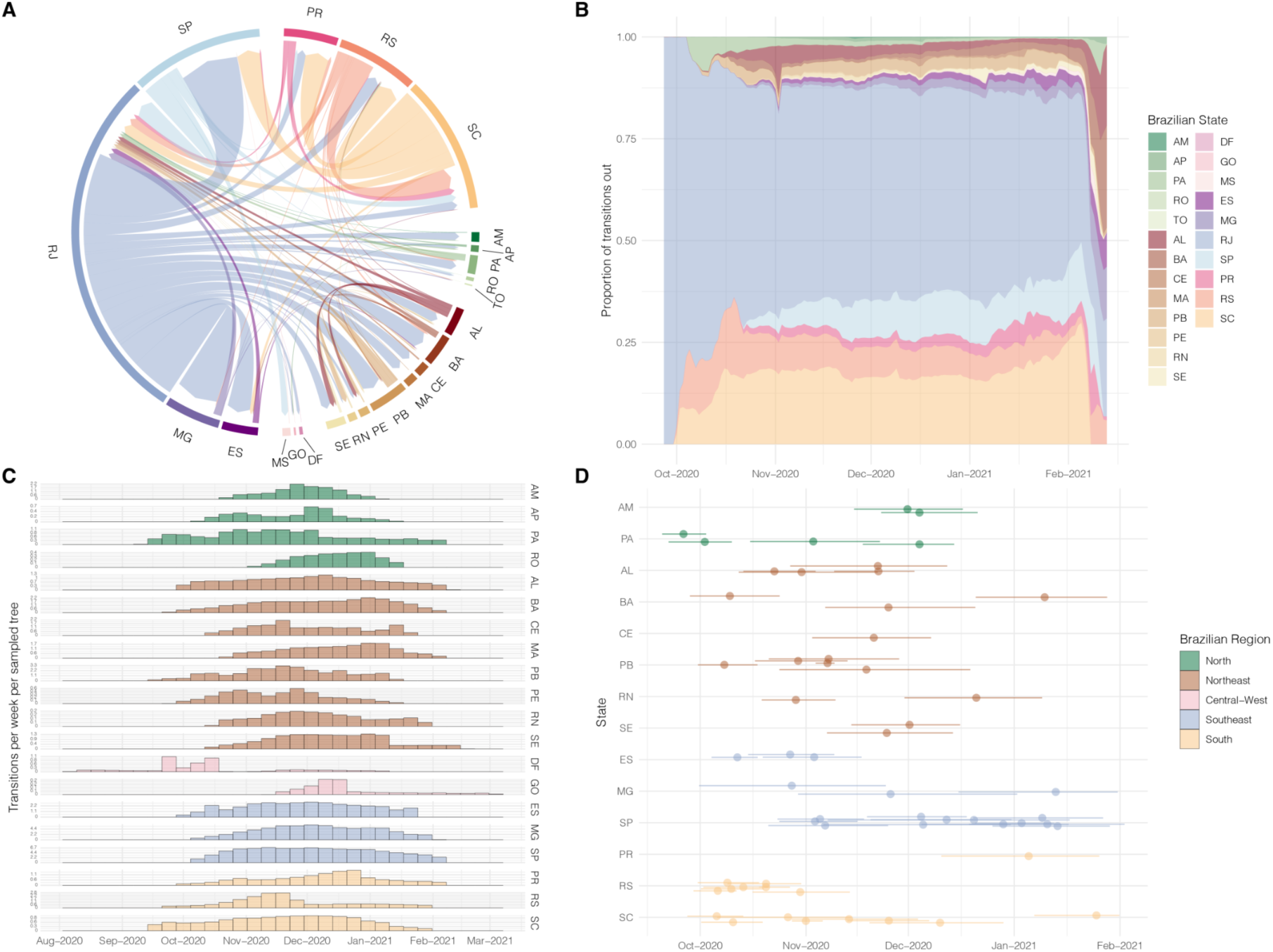
Spatio-temporal spread of SARS-CoV-2 lineage P.2 in Brazil. a) Inter-state circular migration flow plot as estimated from the Markov jumps analysis complementary to the phylogeographic model. Arrows indicate the direction of the migration and thickness is relative to the number of jumps. b) Contribution of each state in seeding P.2 to other locations through-time measured as the proportion of the total Markov per day. c) Time distribution (95% HPD) of jumps from RJ to other Brazilian states. d) T_MRCA_ of each highly supported (SH-aLRT > 80) state specific clades with more than three genomes.

Bayesian analyses support that the VOC Gamma most probably (*PSP* = 1.0) arose in the AM (Northern region) and this state was also the main hub of Gamma spread throughout the country, followed by the Southeastern states of SP and RJ (**Fig 4a and Supplementary Video 2**). AM was responsible for 60-70% of the inter-state disseminations of the VOC Gamma between December 2020 and March 2021, while the three major spreading states combined accounted for >90% of all inter-state viral transmissions in that period (**Fig 4b**). The relative contribution of the secondary hubs (SP and RJ) to the overall dissemination of Gamma in Brazil increased from mid-December 2020 to late March 2021. The first transition events of VOC Gamma from AM into other Brazilian states occurred in December 2020 and reached a peak between January and February 2021 **(Fig 4c**); while the earliest timing of community transmission of this VOC in most Brazilian states was traced to December 2020 **(Fig 4d**). By early January 2021, this variant was already disseminated and established local transmission chains in nine out of 23 Brazilian states from all major country regions: North (Pará), Northeastern (Bahia, Maranhão and Sergipe), Southeastern (SP and RJ), Southern (SC and Paraná) and Central-Western (Goiás) regions.

**Figure 4.**
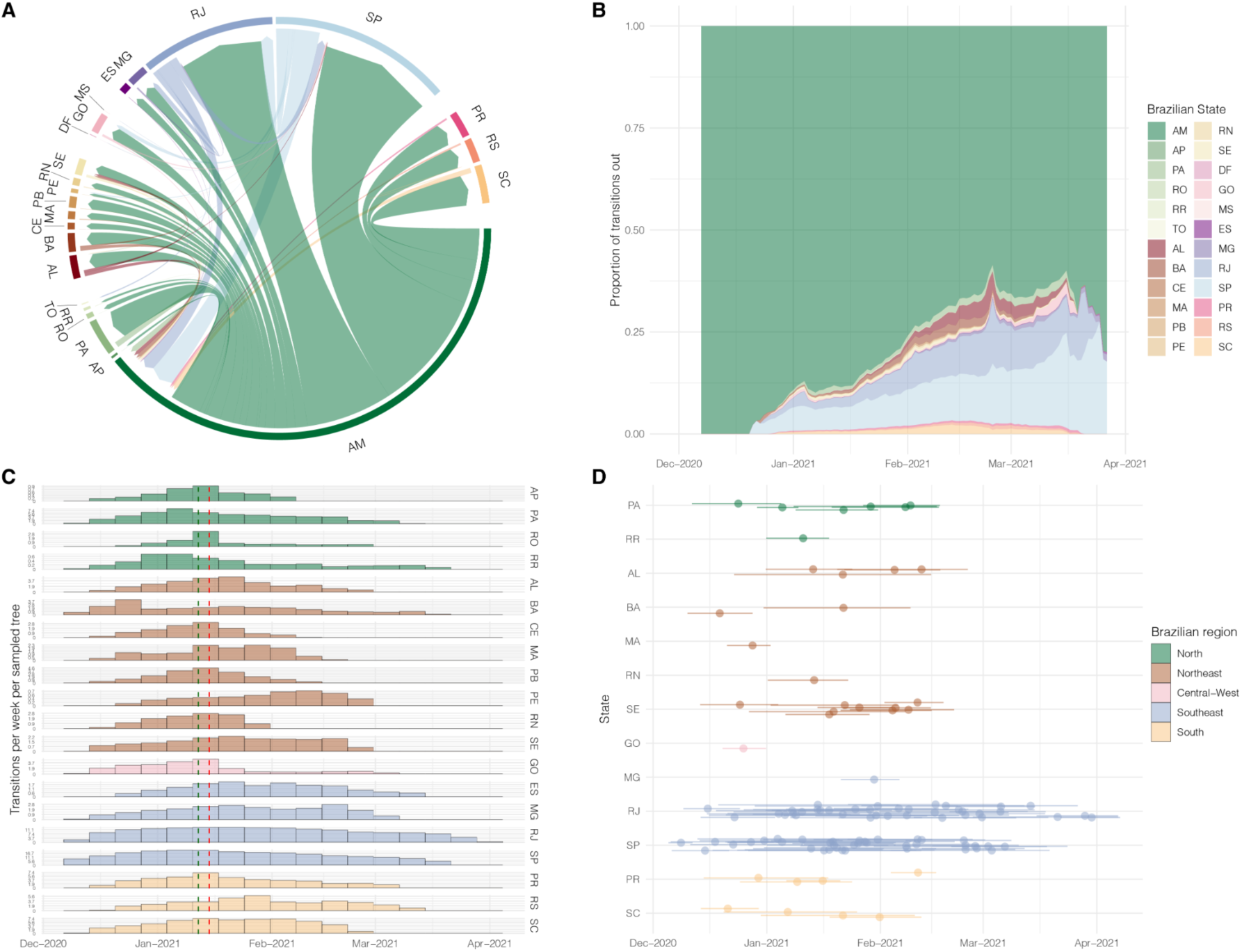
Spatio-temporal spread of SARS-CoV-2 VOC Gamma in Brazil. a) Inter-state circular migration flow plot as estimated from the Markov jumps analysis complementary to the phylogeographic model. Arrows indicate the direction of the migration and thickness is relative to the number of jumps. b) Contribution of each state in seeding Gamma to other locations through-time measured as the proportion of the total Markov jumps per day. c) Time distribution (95% HPD) of jumps from AM to other Brazilian states. The vertical green line shows the time of Gamma first notification and vertical red line shows the starting date of COVID-19 patients transferring from AM to other states. d) T_MRCA_ of each highly supported (SH-aLRT > 80) state specific clades with more than three genomes.

### Estimating the Re of variants Gamma and P.2 in Brazil

We applied the birth-death skyline (BDSKY) model to estimate the Re of P. 2 and Gamma at source locations by selecting all P.2 sequences from RJ (P.2_RJ_) and all Gamma sequences from AM (Gamma_AM_); and at secondary outbreaks by combining large (n > 10) highly supported (SH-aLRT>0.80) clades representative of the circulation in a single state. Only Gamma clusters from SP and RJ, and none for P.2, met the criteria for analysis of secondary outbreaks (see Methods). The temporal trajectory of the median Re of clade P.2_RJ_ supports an initial expansion phase (Re > 1), followed by a phase of stabilization (Re ∼ 1) and subsequent decrease (Re < 1) (**Fig 5a**). Interestingly, when clade P.2_RJ_ reached the highest median Re in September 2020, the lineage P.2 was still undetected ^4^ and the number of SARI cases remained relatively constant in RJ (**Fig 5a**). When lineage P.2 became the dominant variant in RJ in December 2020, the clade P.2_RJ_ displayed a median Re∼1.

**Figure 5.**
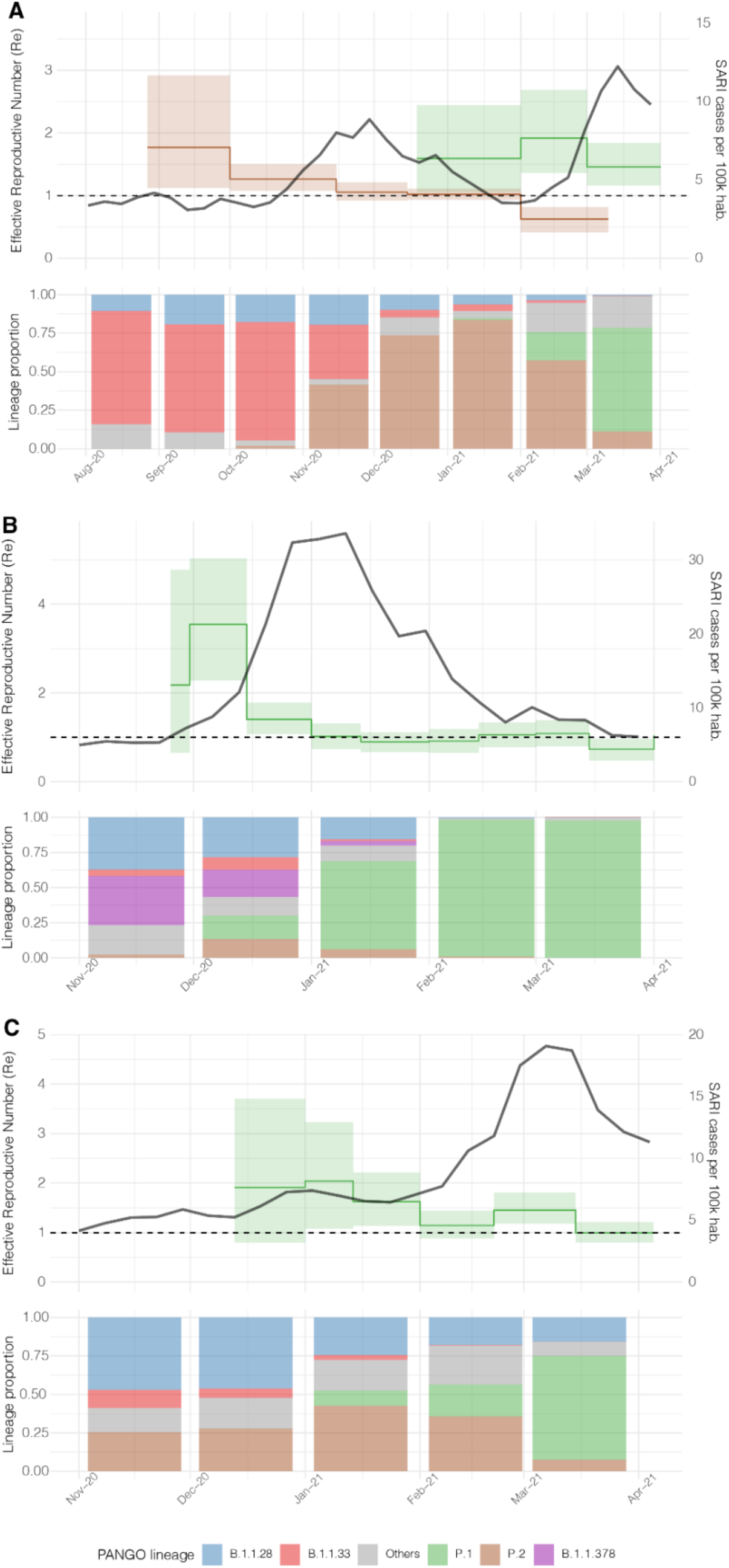
Phylodynamics analyses of variants P.2and Gamma in RJ (a), AM (b) and SP (c) states. Effective reproductive number (Re) [median (solid lines) and 95% HPD (high posterior density)], as estimated by Bayesian phylogenetics methods, is shown in green and brown for Gamma and P.2, respectively. Along with Re, SARI cases are shown as a black line. PANGO lineages proportions per month are shown in the bottom panel, colored according to the legend.

Similarly, to P.2 in RJ, in AM the clade Gamma_AM_ showed an epidemic dynamic with three phases of expansion (Re > 1), stabilization (Re ∼ 1) and subsequent decrease (Re < 1) (**Fig 5b**). When clade Gamma_AM_ reached the highest median Re in December 2020, the relative prevalence of Gamma in Amazonas was low (<10%); but the number of SARI cases already started to increase (**Fig 5b**). When Gamma became the dominant variant in AM in January 2021, the clade Gamma_AM_ also displayed a median Re∼1. The initial expansion phase of major local Gamma clades in RJ (Gamma_RJ_) and SP (Gamma_SP_) from late December 2020 to late January 2021 occurred when Gamma was barely detected, and the number of SARI cases remained relatively constant in those states (**Fig 5a and 5c**). The clade Gamma_RJ_ expanded in RJ during a period (December 2020 to March 2021) when the prevalence of clade P.2_RJ_ was declining and the ratio Re(Gamma_RJ_)/Re(P.2_RJ_) was 1.56 (December-January), 3.06 (February) and 2.33 (March) along the co-circulating period, supporting the higher transmissibility of the VOC Gamma concerning lineage P.2. Comparison of the median Re of VOC Gamma in different states revealed that clade Gamma_AM_ displayed a much faster initial expansion (Re = 3.55) than clades Gamma_RJ_ and Gamma_SP_ (Re = 1.59 and 1.91, respectively). The growth rate of clade Gamma_AM_, however, rapidly decreased in the second half of December 2020 (Re =1.40) and during early 2021 (Re =<1); while clades Gamma_RJ_ and Gamma_SP_ maintained longer phases of epidemic growth (Re = 1.45 - 2.01) until March 2021.

## Discussion

We combine spatial and genomic data to reconstruct the spatiotemporal dynamics of the VOC Gamma and lineage P.2 in Brazil from September 2020 through April 2021, a study period that covers a decreasing phase of COVID-19 incidence from September to November 2020 and a subsequent increasing phase up to March-April 2021. Although both SARS-CoV-2 variants harbor the same mutation of concern S:E484K, they displayed different spatiotemporal dynamics probably shaped by the higher transmissibility of the VOC Gamma concerning lineage P.2.

Our study indicates that variants P.2 and Gamma probably arose in RJ and AM states around July and November 2020, respectively. This is consistent with initial estimations ^4,7,15^ and rejects the hypothesis of a much older origin of lineage P.2 in February 2020 and of VOC Gamma in August 2020. Our analyses suggest that some Gamma and P.2 Brazilian sequences with the earliest recorded sampling times available at the EpiCoV database in GISAID are probably the result of sample contamination or spelling errors in metadata entries. The inclusion of those problematic sequences, as well as of P.1 sequences belonging to Gamma-like clades ^6^, certainly pushed back the T_MRCA_ of Brazilian variants to unreliable old dates. The emergence of lineage P.2 in February 2020 is highly improbable because it overlaps with the estimated origin of the parental lineage B.1.1.28 ^2^, while the emergence of the VOC Gamma in August 2020 is inconsistent with the absence of this variant among hundreds of Amazonian SARS-CoV-2 positive samples genotyped between August and November 2020 ^3^.

Although the densely populated and well-connected states in the Southeastern region (RJ and SP) were the main sources of dissemination of lineage P.2 in Brazil, this variant spread at a slower rate than the VOC Gamma that was mainly disseminated from the poorly connected state of AM. According to our estimations, the earliest onset date of community transmission of lineage P.2 in most sampled states was traced to October 2020 (∼3 months after its emergence), coinciding with the first detection of this variant in such locations^15^, and it only became the most prevalent variant is some states around December 2020 (∼5 months after its emergence). By comparison, the earliest timing of community transmission of the VOC Gamma in many Brazilian states was traced to December 2020 (∼1 month after its emergence) and became the most prevalent variant in all Brazilian states by February 2021 (∼3 months after its emergence). By the time when the VOC Gamma was first described on 12th January 2021 in Japanese travelers returning from the Amazonas state ^16^, this variant had already established local transmission chains in at least nine out of 23 Brazilian states from all major country regions.

The rapid geographic spread of the VOC Gamma and the consistent fixation as the most prevalent SARS-CoV-2 lineage across all Brazilian regions during 2021 supports that this VOC is more transmissible than lineage P.2. The comparison of the Re of local clades of variants Gamma and P.2 that co-circulated in the RJ state from December 2020 to March 2021 allowed us to quantify the relative transmissibility of those lineages while controlling for confounding ecological factors such as local levels of immunity and social distancing measures. Our phylodynamics estimates indicate that the VOC Gamma displayed a median Re that was between 1.56 and 3.06 higher than that of lineage P.2 during the period of co-circulation. Notably, this result is consistent with a previous study conducted in the AM state that estimated that the median Re of the VOC Gamma was 2.2 times higher than that estimated for local B.1.1.28 clades that co-circulated during December 2020 ^3^. These findings confirm the notion that the VOC Gamma was more transmissible than pre-existing SARS-CoV-2 lineages in Brazil, including the lineage P.2, in different local settings.

Even though our phylogeographic reconstructions support that RJ and SP were important hubs of inter-regional spread of SARS-CoV-2 during the epidemic wave in late 2020 and early 2021 as observed in early 2020 ^2^, they also pointed those other Brazilian states from the Northern (AM) and Southern (SC and RS) regions accounted for a substantial fraction of all inter-state disseminations of variants Gamma and P.2, respectively. Our findings also support that inter-state viral transmissions not only occurred between close geographic locations, but also between distant states of Northern, Southern and Northeastern regions. The diversification of geographic hubs of viral dispersion and the detection of long-distance viral migrations suggests that reactivation of domestic passenger air traffic since the second half of 2020 may have been an important driver of dissemination of variants P.2 and Gamma in Brazil. These findings also emphasize that implementation of more strict control measures for domestic air passengers, like the mandatory exigence of a negative SARS-CoV-2 RT-PCR test, could be an effective measure to reduce the dissemination rate of SARS-CoV-2 in Brazil ^17^.

Our findings revealed that variants P.2 and Gamma were disseminated for several weeks before being detected in a new location and their early phase of cryptic expansion is challenging to detect or predict by epidemiological surveillance systems. When the lineage P.2 reached the highest median Re in September 2020 in RJ, the mean daily number of SARS-CoV-2 cases in the state was decreasing and the incidence of SARI cases only started to increase in mid-October 2020. Similarly, the VOC Gamma in RJ and SP probably started to expand in December 2020, well-before the first detection of this variant in January 2021 and the peak of SARI cases associated with Gamma in March 2021 in both states. We hypothesize that the unnoticed early expansion of variants P.2 or Gamma in many locations happened in an epidemic scenario where previous dominant lineages were declining, so the growth of daily SARI cases only became apparent when these new variants were already established as the most prevalent lineage. It may also be possible that initial circulation of variants P.2 and Gamma might have been driven by young people and SARI cases only increased when older individuals got infected.

Because the levels of immunity and social distancing varied over time among different Brazilian states, we compared the Re of the VOC Gamma in AM, RJ and SP between December 2020 and March 2021 to quantify the potential impact of ecological factors on Gamma spread. Although the Re estimates displayed large credibility intervals with considerable overlap, our data suggests that the VOC Gamma displayed a much faster initial expansion in AM (Re ∼ 3.6) than in RJ or SP (Re ∼ 1.5-2.0), but the growth phase in AM extends over a shorter time than in the Southeastern states. We speculate that the weaker mitigation measures implemented in AM with respect to RJ and SP at late 2020 and/or large-scale gatherings may have amplified the intrinsic higher transmissibility of the VOC Gamma in this Northern state ^3^. The steep decrease of Re in AM in January 2021 might be the result of non-pharmaceutical interventions (NPIs) implemented after the health system collapse^3^ and/or of high levels of population immunity estimated at ∼70% in Manaus^18^. On the other hand, the larger number of susceptible individuals in the Southeastern region may have sustained a longer phase of Gamma expansion.

The most important limitation of our study was the uneven sampling among Brazilian states and throughout the time, which may have introduced some bias in phylogeographic analyses. Although the number of genomes analyzed in this study roughly follows the number of COVID-19 confirmed cases in Brazil (**Figure 1A and 1B**), the smaller sampling from September and December 2020 limited the potential of our analyzes to identify large state-specific P.2 clusters and to accurate estimates the earliest onset date of communitarian transmission of this variant in several locations. In addition, even though the datasets here analyzed comprise SARS-CoV-2 sequences from 24 out of 27 Brazilian states, the uneven distribution of these genomes among locations might have biased contributions to the overall inter-states’ transmissions.

In summary, this molecular epidemiological study showed that the COVID-19 epidemic in Brazil from September 2020 to March 2021 was characterized by the emergence and spread of the lineage P.2 and the VOC Gamma that were the most prevalent SARS-CoV-2 variants at different time periods in the country. The spatial dispersion of these variants in Brazil was driven by short and long-distance viral transmission and both variants circulated cryptically in several locations for some weeks before being detected. The VOC Gamma displayed a higher transmissibility than lineage P.2 which explained the faster rate of spatial spread of this variant and its establishment as the dominant SARS-CoV-2 lineage in Brazil within a few (∼3) months after its emergence. Our findings also support that the transmissibility of the VOC Gamma varied according to the geographic context probably due to regional differences in social distancing measures and/or fraction of population previously infected. These findings also strengthen the need for ongoing genomic surveillance to provide early warnings about spread of emergent SARS-CoV-2 variants in Brazil.

## Material and Methods

### The genomic surveillance network and ethical aspects

The Brazilian Ministry of Health COVID-19 Genomic Surveillance Network is composed by Oswaldo Cruz Foundation (Fiocruz), Adolfo Lutz Institute (IAL), Evandro Chagas Institute (IEC), the Central Laboratories of each State of Brazil (LACENs) and other partners, such as Health Secretariat of Aparecida de Goiânia, Goiás. Saliva or nasopharyngeal swabs (NPS) samples were collected from suspect COVID-19 cases by sentinel hospitals or health care units and initially tested by real time RT-PCR as a routine diagnostic for SARS-CoV-2 by LACENs using any of the following different commercial assays: SARS-CoV2 (E/RP) (Biomanguinhos), Allplex 2019-nCoV Assay (Seegene) or an in-house protocol following the USA/CDC guidelines (https://www.fda.gov/media/134922/download). SARS-CoV-2 positive samples with cycling threshold (Ct) below 25 were randomly selected for genome sequencing. Alternatively, targeted cases, such as reinfection cases and fatal cases without comorbidities, were also selected for sequencing. Samples are then sent to one of the genomic reference laboratories participating in the network for whole genome sequencing. This study was approved by the Ethics Committee of the of the FIOCRUZ (CAAE: 68118417.6.0000.5248) and Amazonas State University (CAAE: 25430719.6.0000.5016), which waived the signed informed consent.

### SARS-CoV-2 amplification and sequencing

The viral RNA was subjected to reverse transcription and PCR amplification using in-house protocols developed by COVID-19 Fiocruz Genomic Network ^19,20^ or the Illumina COVIDSeq Test (Illumina), including some primers to cover regions with dropout ^14^. Normalized pooled amplicons of each sample were used to prepare NGS libraries with Nextera XT (FC-131-1096) and clustered with MiSeq Reagent Kit v2 (500-cycles - MS-102-2003) on 2 × 250 cycles (in-house protocols) or 2 × 150 cycles (MS-103-1002) paired-end runs. All sequencing data was collected using the Illumina MiSeq sequencing platforms and MiSeq Control software v2.6.2.1 (Illumina).

### SARS-CoV-2 whole-genome consensus sequences and genotyping

FASTQ reads were generated by the Illumina pipeline at BaseSpace (https://basespace.illumina.com). All files were downloaded and imported into Geneious v10.2.6 for trimming and assembling using a customized workflow employing BBDuk and BBMap tools (v37.25) and the NC_045512.2 RefSeq as a template. Using a threshold of at least 50% to call a base. Consensus sequences were initially assigned to viral lineages according to the nomenclature proposed by Rambaut et al. ^21^, using the Pango Lineage software (https://pangolin.cog-uk.io) and later confirmed using phylogenetic analyses.

### Dataset assembling

We complemented the P.1 and P.2 genomes dataset identified in our genomic surveillance by retrieving all Brazilian high quality (<5% N) whole-genomes (29 kb) identified as P.1 and P.2 from EpiCoV database in GISAID (https://www.gisaid.org) and that were deposited up to April 16th. Mutational composition of these sequences was analyzed in NextClade (https://clades.nextstrain.org) and due to highly variable patterns of substitutions, we have retained in the dataset only those genomes with the full-set of P.2 and Gamma lineages defining synapomorphies, thus excluding from the analysis all Gamma-like sequences (**Supplementary Table 2 and 3**).

### Analysis of temporal signal

SARS-CoV-2 Gamma and P.2 complete genome sequences were aligned using MAFFT v7.467 ^21^ and subject to maximum likelihood (ML) phylogenetic analysis using IQ-TREE v2.1.2 ^22^ under the general time-reversible (GTR) model of nucleotide substitution with a gamma-distributed rate variation among sites, four rate categories (G4), a proportion of invariable sites (I) and empirical base frequencies (F) nucleotide substitution model, as selected by the ModelFinder application ^23^. The branch support was assessed by the approximate likelihood-ratio test based on the Shimodaira– Hasegawa-like procedure (SH-aLRT) with 1,000 replicates. The temporal signal of the P.1 and P.2 assembled datasets was assessed from the ML tree by performing a regression analysis of the root-to-tip divergence against sampling time using TempEst ^24^ and excluding outlier sequences that deviate more than 1.5 interquartile ranges from root-to-tip regression line, which included those Gamma and P.2 sequences with the oldest sampling dates. The expected collection dates of the oldest Gamma and P.2 sequences were next estimated by using the Bayesian leaf-dating method developed to estimate the age of sequences with unknown collection date ^25^. For this analysis, the oldest Gamma and P.2 sequences were combined with sequences from Amazonas (Gamma epicenter) and RJ P.2 epicenter), respectively, and were analyzed with the BEAST v1.10 software ^26^ using a GTR+F+I+G4 nucleotide substitution model, an exponential growth coalescent model for the tree prior and a strict molecular clock with a uniform substitution rate prior (8 - 10 × 10^−4^ substitutions/site/year). To estimate the expected sampling dates of the oldest sequences we set a precision value of two years for the recorded date (year and month) of sampling and select the tip date option sampling uniformly from precision. Sequences for which the recorded collection date was not contained within the 95% High Posterior Density (HPD) interval of the leaf-age estimate were excluded. Markov Chain Monte Carlo (MCMC) was run sufficiently long to ensure convergence (effective sample size> 200) in all parameter estimates as assessed in TRACER v1.7 ^27^.

### Estimating time-scaled phylogenetic trees

Due to the size of the datasets, we firstly estimated the time of the most recent common ancestor (T_MRCA_) of each lineage by applying a full phylogenetic Bayesian analysis in downsampled datasets. To do so, 10 genomes per Brazilian state per week were randomly chosen using Augur ^28^, resulting in a dataset of 191 Gamma and 219 P.2 sequences. Alignments were generated using MAFFT v7.475 ^21^ and visually inspected in AliView ^29^. Time-scaled phylogenetic trees were estimated in BEAST v.1.10 ^26^ under a GTR+F+I+G4 model. The evolutionary model was complemented with the non-parametric Bayesian skyline (BSKL) model as the coalescent tree prior ^30^ and a strict molecular clock model with a uniform substitution rate prior (8 - 10 × 10^−4^ substitutions/site/year). MCMC was run sufficiently long to ensure convergence (effective sample size> 200) in all parameter estimates as assessed in TRACER v1.7 ^27^. The maximum clade credibility (MCC) tree was summarized with TreeAnnotator v1.10. ML and MCC trees were visualized using FigTree v1.4.4 (http://tree.bio.ed.ac.uk/software/figtree/). The time-scaled tree for the full Gamma (N=2,477) and P.2 (N=1,357) datasets was performed using a modified version of BEAST (https://beast.community/thorney_beast) that makes feasible the analysis of big datasets. In summary the method alleviates most of the computational burden by fixing the tree topology and then re-scales the branch lengths based on a clock and coalescent models. To do so, ML phylogenetic trees constructed for both Gamma and P.2 datasets as explained above, were inputted in the BEAST xml file as starting and data trees and analyses were performed under a logistic coalescent prior, which outperformed [Bayes Factor (BF) > 3] the exponential prior in a Marginal Likelihood Estimation (MLE) of model fitness, and a strict molecular clock, as specified above. Additionally, we set a uniform prior on the root height estimates based on the 95% HPD determined in the BEAST analysis of the downsampled datasets. Four and three MCMC were run for 100 million generations and then combined to ensure stationarity and good mixing for the Gamma and P.2 datasets, respectively.

### Discrete Bayesian phylogeography

A set of 1000 and 900 trees was randomly selected from the posterior distribution of trees resulting from the BEAST analysis of the full Gamma and P.2 datasets, respectively. Sampling locations (Brazilian states) were used as traits in the phylogeographic model and reconstruction of the ancestral states was performed using a discrete symmetric model with BSSVS ^31^ on the posterior sampling of trees. SPREAD software ^32^ was used to identify the well-supported transition rates based (BF > 3). We complemented this analysis with Markov jump estimation of the number of location transitions throughout evolutionary history ^33^, which was used to explore directionality of the transitions. Viral migration history between all supported location transitions was then visualized in circular migration flow plots using the package “circlize” ^34^ available in R software (https://www.r-project.org). The temporal history of transitions from the lineage epicenter to other Brazilian states was summarized as histograms in one-week periods. Because many spatial transitions connected nodes with low support, we used a conservative approach to estimate the onset date of community transmission in each state from the T_MRCA_ of only highly supported (SH-aLRT > 80) clusters comprising at least three sequences from a given state and whose location root was most probably traced (*Posterior State Probability* [*PSP*] > 0.50) to that state. Additionally, we summarize P.2 and Gamma lineages spatial-temporal history alongside with SARI cases trajectories in Brazil, by generating an animation with baltic library (https://github.com/evogytis/baltic).

### Effective Reproductive Number (Re) Estimation

To estimate the Re of lineages Gamma and P.2 through time in the source locations, we selected all Gamma sequences from Amazonas and all P.2 sequences from RJ whose state of the ancestral node in the phylogeographic trees remained in Amazonas or RJ, respectively. To estimate the Re of Gamma and P.2 dissemination outside the source locations, state specific clusters in the phylogenetic trees were selected based on the following criteria: a) high branch support (SH-aLRT > 80); b) cluster size bigger than 10 sequences; c) > 80% of sequences from a single state and origin of the cluster in the same state. Selected clusters were then combined and analyzed when the dataset comprised more than 50 sequences. Following these criteria, we could only analyze the phylodynamic of local dissemination of Gamma in the states of RJ and SP. The assembled datasets comprised 298 and 199 Gamma genomes for RJ and SP, respectively. For P.2 we could not analyze local dissemination outside the source region. The assembled datasets were then analyzed using the birth-death skyline (BDSKY) model ^35^ which reconstructs Re trajectories and is implemented within BEAST 2 v2.6.5 ^36^. The sampling rate (d) was set to zero for the period before the oldest sample and estimated from the data afterward. The BDSKY prior settings were as follows: Become Uninfectious Rate (exponential, mean = 36); Reproductive Number (log normal, mean = 0.8, sd = 0.5); Sampling Proportion (beta, alpha = 1, beta = 100). Origin parameter was conditioned to the root height, and the Re was estimated in a piecewise manner over intervals defining to match state specific epidemiological trajectories. Molecular clock was as in the time-scaled trees analysis and the HKY+G4+F substitution model was used. MCMC chains were run until all relevant parameters reached ESS > 200, as explained above.

## Supporting information

Supplementary Table 4 - GISAID acknowledgement

## Data Availability

All data produced are available online at the EpiCoV database in GISAID (https://www.gisaid.org/)

## Acknowledgements

The authors wish to thank all the health care workers and scientists who have worked hard to deal with this pandemic threat, the GISAID team, and all the EpiCoV database’s submitters. GISAID acknowledgment table containing sequences used in this study is shown in **Supplementary Table 4**. We also appreciate the support of the Fiocruz COVID-19 Genomic Surveillance Network (http://www.genomahcov.fiocruz.br/) members, the Respiratory Viruses Genomic Surveillance Network of the General Laboratory Coordination (CGLab), Brazilian Ministry of Health (MoH), Brazilian Central Laboratory States (LACENs), and the Amazonas surveillance teams for the partnership in the viral surveillance in Brazil.

## Appendix 1

**Authors members of Brazilian Ministry of Health COVID-19 Genomic Surveillance Network**

Tirza Peixoto Mattos^1^, Valdinete Alves Nascimento^2^, Victor Souza^2^, André de Lima Guerra Corado^2^, Fernanda Nascimento^2^, George Silva^2^, Matilde Mejía^2^, Maria Júlia Brandão^2^, Ágatha Costa^2^, Karina Pessoa^2^, Michele Jesus^2^, Luciana Fé Gonçalves^2^, Cristiano Fernandes^3^, Valnete Andrade^4^, Luana Barbagelata^5^, Ana Cecília Ribeiro Cruz^5^, Andrea Costa^6^, Lindomar dos Anjos Silva^6^, Jucimária Dantas Galvão^7^, Anderson Brandao Leite^8^, Felicidade Mota Pereira^9^, Thais Oliveira Costa^10^, Joaquim Cesar Sousa Jr^10^, Lidio Gonçalves Lima Neto^11^, Haline Barroso^12^, Dalane Loudal Florentino Teixeira^12^, Joao Felipe Bezerra^13^, Cássia Docena^14^, Raul Emídio de Lima^14^, Lilian Caroliny Amorim Silva^14^, Gustavo Barbosa de Lima^14^, Laís Ceschini Machado^14^, Matheus Filgueira Bezerra ^14^, Marcelo Henrique Santos Paiva^14,15^, Maria Eduarda Pessoa Lopes Dantas^16^, Raíssa Liane Do Nascimento Pereira^16^, Josélio Araújo^16^, Cliomar A Santos^17^, Rodrigo Ribeiro Rodrigues^18^, André Felipe Leal Bernardes^19^, Felipe Campos de Melo Iani^19^, Beatriz Grinsztejn^20^, Valdiléa G Veloso^20^, Patricia Brasil^20^, Anna Carolina Dias da Paixão^21^, Luciana Reis Appolinario^21^, Renata Serrano Lopes^21^, Fernando do Couto Motta^21^, Alice Sampaio Rocha^21^, Taina Moreira Martins Venas^21^, Elisa Cavalcante Pereira^21^, Andrea Cony Cavalcanti^22^, Leonardo Soares Bastos^23^, Luis Fernando de Macedo Brigido^24^, Mauro de Medeiros Oliveira^25^, Michelle Orane Schemberger^25^, Andreia Akemi Suzukawa^25^, Irina Riediger^26^, Maria do Carmo Debur^26^, Richard Steiner Salvato^27^, Tatiana Schäffer Gregianini^27^, Darcita Buerger Rovaris^28^, Sandra Bianchini Fernandes^28^

**Affiliations**

^1^Laboratório Central de Saúde Pública do Amazonas (LACEN-AM), Manaus, Amazonas, Brazil; ^2^Laboratório de Ecologia de Doenças Transmissíveis na Amazônia (EDTA), Leônidas e Maria Deane Institute, Fiocruz; ^3^Fundação de Vigilância em Saúde do Amazonas - Dra. Rosemary Costa Pinto, Manaus, AM, Brazil; ^4^Laboratório Central de Saúde Pública do Pará (LACEN-PA), Belém, Pará, Brazil; ^5^Instituto Evandro Chagas, Belém, Pará, Brazil; ^6^Laboratório Central de Saúde Pública do Amapá (LACEN-AP), Macapá, Amapá, Brazil; ^7^Laboratório Central de Saúde Pública do Tocantins (LACEN-TO), Palmas, Tocantins, Brazil; ^8^Laboratório Central de Saúde Pública de Alagoas (LACEN-AL), Maceió, Alagoas, Brazil; ^9^Laboratório Central de Saúde Pública Prof. Gonzalo Moniz (LACEN-BA), Salvador, Bahia, Brazil; ^10^Fundação Oswaldo Cruz - Fiocruz Ceará; ^11^Laboratório Central de Saúde Pública do Maranhão (LACEN-MA), São Luís, Maranhã, Brazil; ^12^Laboratório Central de Saúde Pública da Paraíba (LACEN-PB), João Pessoa, Paraíba, Brazil; ^13^Universidade Federal da Paraíba (UFPB); ^14^Instituto Aggeu Magalhães, Fundação Oswaldo Cruz, Recife, Pernambuco, Brazil; ^15^Núcleo de Ciências da Vida, Centro Acadêmico do Agreste, Universidade Federal de Pernambuco (UFPE), Caruaru, Pernambuco, Brazil; ^16^Universidade Federal do Rio Grande do Norte (UFRN); ^17^Laboratório Central de Saúde Pública de Sergipe (LACEN-SE), Aracaju, Sergipe, Brazil; ^18^Laboratório Central de Saúde Pública do Espirito Santo (LACEN-ES), Vitória, Espirito Santo, Brazil; ^19^Laboratório Central de Saúde Pública de Minas Gerais (LACEN-MG), Belo Horizonte, Minas Gerais, Brazil; ^20^Instituto Nacional de Infectologia Evandro Chagas (INI), Fiocruz, Rio de Janeiro, Brazil; ^21^Laboratory of Respiratory Viruses and Measles, Oswaldo Cruz Institute (IOC), Oswaldo Cruz Foundation (FIOCRUZ), Rio de Janeiro, RJ, Brazil; ^22^Laboratório Central de Saúde Pública do Rio de Janeiro (LACEN-RJ), Rio de Janeiro, Rio de Janeiro, Brazil; ^23^Grupo de Métodos Analíticos em Vigilância Epidemiológica, Programa de Computação Científica (PROCC), Fiocruz, Rio de Janeiro, Brazil; ^24^Instituto Adolfo Lutz, São Paulo; ^25^Instituto Carlos Chagas (ICC), Fiocruz-PR; ^26^Laboratório Central de Saúde Pública do Paraná (LACEN-PR), Curitiba, Paraná, Brazil; ^27^Laboratório Central de Saúde Pública do Rio Grande do Sul (LACEN-RS), Porto Alegre, Rio Grande do Sul, Brazil; ^28^Laboratório Central de Saúde Pública do Estado de Santa Catarina (LACEN-SC), Florianópolis, Santa Catarina, Brazil.

