## Supplementary Material for "Phylogenetic-based inference reveals distinct transmission dynamics of SARS-CoV-2 variant of concern Gamma and lineage P.2 in Brazil"

**Supplementary Table 1.** Genomes sampling spatial distribution

| State | Generated in this study | Available in GISAID | Total analyzed |
|---|---|---|---|
| AC | 0 | 98 | 98 |
| AL | 152 | 40 | 192 |
| AM | 616 | 215 | 831 |
| AP | 30 | 46 | 76 |
| BA | 173 | 148 | 321 |
| CE | 112 | 14 | 126 |
| DF | 16 | 7 | 23 |
| ES | 132 | 47 | 179 |
| GO | 607 | 135 | 742 |
| MA | 111 | 34 | 145 |
| MG | 155 | 191 | 346 |
| MS | 71 | 11 | 82 |
| MT | 11 | 13 | 24 |
| PA | 184 | 41 | 225 |
| PB | 103 | 87 | 190 |
| PE | 41 | 7 | 48 |
| PI | 1 | 17 | 18 |
| PR | 295 | 181 | 476 |
| RJ | 552 | 1034 | 1586 |
| RN | 73 | 81 | 154 |
| RO | 25 | 1 | 26 |
| RR | 16 | 16 | 32 |
| RS | 175 | 459 | 634 |
| SC | 392 | 41 | 433 |
| SE | 228 | 46 | 274 |
| SP | 1135 | 3228 | 4363 |
| TO | 66 | 6 | 72 |
| Not available | 0 | 8 | 8 |

Brazilian states' names follow the International Organization for Standardization (ISO) 3166-2 standard

**Supplementary Table 2.** Gamma lineage defining synapomorphies

| Gene | Amino acid | Nucleotide |
|---|---|---|
| ORF1a (NSP1) | - | T733C |
| ORF1a (NSP3) | - | C2749T |
| ORF1a (NSP3) | S1188L (S370L) | C3828T |
| ORF1a (NSP3) | K1795Q (K977Q) | A5648C |
| ORF1a (NSP3) | - | A6319G |
| ORF1a (NSP3) | - | A6613G |
| ORF1a (NSP6) | - | del: 11288-11296 |
| ORF1a (NSP9) | - | C12778T |
| ORF1b (NSP12) | - | C13860T |
| ORF1b (NSP13) | E1264D (E341D) | G17259T |
| S | L18F | C21614T |
| S | T20N | C21621A |
| S | P26S | C21638T |
| S | D138Y | G21974T |
| S | R190S | G22132T |
| S | K417T | A22812C |
| S | E484K | G23012A |
| S | N501Y | A23063T |
| S | H655Y | C23525T |
| S | T1027I | C24642T |
| ORF3a | S253P | T26149C |
| ORF8 | E92K | G28167A |
| N/ORF9b | P80R/Q77E | C28512G |
| ORF8/N intergenic | - | ins: 28269 - 28273 |

**Supplementary Table 3.** P.2 lineage defining synapomorphies

| Gene | Amino acid | Nucleotide |
|---|---|---|
| 5`UTR | - | C100T |
| ORF1a (Mpro) | L3468V | T10667G |
| ORF1a (NSP6) | - | C11824T |
| ORF1a (NSP7) | L3930F | C12053T |
| S | E484K | G23012A |
| ORF8 | - | C28253T |
| N | A119S | G28628T |

**Supplementary Video 1 and 2**

Map of Brazil with different colors for each state is shown on the left. Inferred movements of SARS-CoV-2 P.2 lineage (Video 1) and VOC Gamma (Video 2) are indicated with tapered arrows, colored by its origin state. Phylogenetic tree in the upper right shows virus migration history as reconstructed by the phylogeographical analysis, with branches colored by location (Brazilian states). Migrations inferred between any two locations in the tree are animated on the map on the left. Plot on the lower right shows the sum of weekly SARI cases per 100k inhabitants reported for each Brazilian state. SARI cases are the sum of any SARS-CoV-2 lineages circulating in each week; thus, they do not specifically represent P.2 or VOC Gamma circulation in Video 1 and 2, respectively.
